# Field-Based Assessment of Lymphatic Filariasis Transmission Dynamics and Intervention Outcomes in Varanasi District

**DOI:** 10.1101/2025.07.14.25331498

**Authors:** Shivani Sharma, Amit Kumar Singh, Tripti Singh, Sunil Kumar, Anchal Singh

## Abstract

Background-Lymphatic filariasis is a neglected tropical disease that imposes a significant burden on the mental, physical, and socioeconomic health of millions of people worldwide. India bears a considerable amount of this burden, with LF endemic in 21 states and union territories. The government of India implemented nationwide Mass Drug Administration and Triple Drug Therapy in a few districts, including Varanasi, as a part of the Global Programme to Eliminate Lymphatic Filariasis (GPELF).

Objective: This study intends to assess the IDA coverage in Varanasi from 2019 to 2021 and its effect on LF transmission with the help of night blood tests in different sentinels and spot check sites and to check the microfilariae rate (<1%) with the conduction of TAS in 2022.

Method: In this conducted study regions from both urban and rural consisting of various PHC/CHC in Varanasi district were selected for the MDA/IDA, data were collected annually, Night blood tests were performed following WHO protocols. Microfilariae rates were calculated in the areas and TAS were performed in sentinels and spot check sites to access the ongoing transmission.

Result: The coverage of IDA has significantly improved from 74.79% in 2019 to over 80% in 2020 and 2021. There was a decrease in microfilariae rate observed in sentinel sites, which was 0.94% in 2019 to 0.15% in 2021. Similarly, MF rates declined when checked in sentinel sites, from 0.46% in 2019 to 0.10% in 2021. TAS 2022, which involved 19,852 participants, showed an overall MF rate of 0.1% in most regions. However, Cholapur failed the TAS with a reported MF rate of 1.34%, which has led to more IDA rounds to follow.

Conclusion: This paper highlights the efficacy of IDA in reducing the LF transmission in Varanasi. There were areas that demonstrated substantial progress, while targeted efforts are still required in high-prevalence regions. Ongoing surveillance and strategic intervention are still required to achieve and sustain LF elimination goals till 2027.

## Introduction

Lymphatic filariasis, commonly referred to as Elephantiasis, is a neglected tropical disease that significantly hinders individuals’ ability to engage in work, limits their access to essential services, and contributes to the social stigma faced by those affected and their families. (WHO 2010) The clinical manifestations of the infection can profoundly affect an individual’s mental health, especially in cases where access to essential care for the associated morbidity and disabilities is lacking, resulting in a total of 5.09 million years of disability-adjusted life years. Many infected individuals do not exhibit any symptoms; however, if treatment is not received over a prolonged period, the infection may lead to limb lymphedema in both genders and hydrocele in men. (Ton, Mackenzie et al. 2015, de Souza, Gass et al. 2020) The affected individual and his or her family may suffer severely devastating social and economic repercussions because of these clinical manifestations. Following mental illness, LF is considered the second most debilitating condition globally.(Wynd, Melrose et al. 2007)

WHO’s Global Programme to Eliminate Lymphatic Filariasis (GPELF) was established in 2000 WHO’s approach is built around two main elements: reducing the suffering caused by lymphatic filariasis by providing the suggested necessary package of care, and preventing the spread of infection by treating all eligible individuals on a broad scale each year in an area or region where infection is present. Eighty-one countries were thought to be endemic for lymphatic filariasis at the beginning of the GPELF. Since then, additional epidemiological data reviews have shown that in ten countries, preventive chemotherapy was not necessary. In the year 2023, a total of 657 million individuals in 39 countries lived in areas that required preventive measures to reduce the spread of infection. The global baseline estimate of individuals affected by lymphatic filariasis comprises 25 million males with hydrocele and over 15 million individuals with lymphoedema. At least 36 million individuals persist in experiencing these symptoms associated with chronic illnesses. Eliminating lymphatic filariasis has the potential to reduce unnecessary suffering and contribute to poverty alleviation (https://www.who.int/news-room/fact-sheets/detail/lymphatic-filariasis) India had a large prevalence of LF, with the disease being endemic in 21 states and union territories. Numerous states, including Uttar Pradesh, Bihar, Orissa, Jharkhand, and Andhra Pradesh, have highly endemic areas. Both rural and urban regions are affected by the disease. Although the National Filaria Control Programme (NFCP) was established in 1955, its reach was constrained by resource shortages and operational issues. Population growth and a lack of control measures in many rural and urban regions cause the size of endemic populations and the number of infected individuals to rise dramatically. (https://ncvbdc.mohfw.gov.in/WriteReadData/l892s/1031567531528881007.pdf)

In 2004, the Government of India initiated home-based morbidity management and a nationwide mass drug administration (MDA) in endemic regions to meet established targets, resulting in an increase in the number of hydrocelectomies conducted in hospitals and community health centers (CHCs). In 2004, only 202 districts managed to attain a coverage percentage of 72.6 percent. In 2007, there was an increase in the number of districts, resulting in all 250 recognized LF endemic districts being placed under MDA, which has now risen to 256 due to bifurcation. A total of 257 districts are encompassed within the MDA framework, with one district added in 2019. Collectively, these districts are home to around 650 million individuals who are at risk of lymphatic filariasis across the nation. Approximately 500 million individuals meet the criteria for MDA from the overall population. The honourable Union Health minister and the Minister of State presented the Accelerated Plan, featuring Triple Drug Therapy (IDA), during the Global Alliance Elimination of Lymphatic Filariasis (GAELF) meeting held from June 13 to June 15, 2018. From 2004 to 2019, there was an increase in population coverage during MDA, rising from 73% to 87.33%.In 2019, MDA was recorded across 151 districts. The implementation programme for Triple Drug Therapy (IDA) has been successfully carried out in five districts: Arwal (Bihar), Simdega (Jharkhand), Varanasi (Uttar Pradesh), Nagpur (Maharashtra), and Yadgir (Karnataka), receiving approval from the Ministry of Health and Family Welfare. By December 2019, a total of eleven more districts in Uttar Pradesh had implemented the IDA. In January 2020, the district of Tapi in Gujarat initiated the use of IDA. (govt of India site). The 2002 National Health Policy of India established a target of 2015 for the eradication of LF as a public health issue in the country, which was subsequently extended to 2017. By 2020, the Global Programme to Eliminate Lymphatic Filariasis sought to eradicate the illness. Consequently, the global objective was revised to eliminate LF by 2030. National objectives were achieved in endemic countries to attain this goal. (Tripathi, Roy et al. 2022)

The active participation of healthcare professionals at different levels in the execution of plans and management of morbidity throughout every stage of care is essential for the eradication of lymphatic filariasis. This encompasses health professionals in primary health care and several other tiers of care, along with ASHA at the village level. To guarantee that success is achieved and maintained, supervision, monitoring, and surveillance are essential operational elements for the successful elimination of LF in India.To ensure that success is not only attained but also sustained, supervision, monitoring, and surveillance components are among the most crucial operational components to the success of LF elimination in India. These components also include effective programme management. https://ncvbdc.mohfw.gov.in/WriteReadData/l892s/National-Roadmap-ELF.pdf

WHO recommends four sequential procedures for implementing and targeting MDA are all predicated on clearly established epidemiological indicators. 1.Determine the disease’s geographic distribution through mapping. 2.Monitor baseline infection burden and impact (through sentinel and spot-check sites) prior to the first and fourth rounds of MDA and following the fifth, and assess transmission to determine whether MDA can be stopped. 3.Verify that transmission has been interrupted. 4. Implement MDA annually for five years while reporting drug coverage. (Rebollo and Bockarie 2013)

Several studies have been carried out to evaluate the coverage and compliance of mass medication administration. To evaluate the coverage and adherence to triple medication therapy for lymphatic filariasis in the Varanasi district of Uttar Pradesh, India, the current study was carried out. Triple drug therapy was started in the Varanasi district during the MDA/IDA round 2019–2021. The Transmission Assessment Survey (TAS) is a method used to evaluate the amount of filarial antigen that is circulating in the human population. India had 90% of implementing units covered in 2016. After passing TAS, 94 out of 256 endemic districts no longer continue to administer drugs in large quantities. Lymphatic filariasis usually begins in childhood and progresses into maturity, causing chronic, incurable diseases such hydrocele, lymphoedema, and elephantiasis.(Parveen and SINGH 2023)

## Material and methods

### Ethics statement

The study was accepted by Banaras Hindu University’s Institutional Ethical Committee in Varanasi (Ref No: I.Sc./ECM-XIX/2025-26/ dated: 19.04.2025). After a thorough explanation of the experimental purpose in the local language, written informed consent was obtained from each participant. Prior the trial started; each participant was asked about their disease status and received a physical and clinical assessment.

### Study area and population

This study, which examined several Primary Health Centers (PHCs) and Community Health Centers (CHCs), was conducted in Varanasi, India. The study looked at both urban and rural areas, assessing how the Triple drug therapy (IDA) affected the spread of lymphatic filariasis between 2019 and 2021 that included Night blood tests of the participants. In 2022, a Transmission Assessment Survey (TAS) was conducted for check upon the successful conduction of the IDA.

### Study setting

With an estimated 41 lakh population, the Varanasi District in Uttar Pradesh is made up of 90 wards, 1,360 villages, and four sub-districts spread across 1,535 km². As part of the pre-IDA activities, night blood surveys were conducted on November 15, 16, and 17, 2018, to assess baseline microfilaremia levels prior to intervention The initial phase of Varanasi’s IDA: In 2019, the implementation of IDA aimed to speed up the elimination of LF within the district. The initiative commenced on February 20, 2019, and continued until March 2, 2019, addressing the substantial population across both rural and urban regions of the district. This was promptly succeeded by additional rounds aimed at reaching all remaining beneficiaries, especially within anganwadis, schools, colleges, government, and private offices, as well as other relevant areas. The Anganwadi centre serves as a government initiative within the village, focusing on nutrition, health screening, and health education for children aged 0–6 years, as well as for adolescent girls, pregnant women, and lactating mothers.

Individuals at the village level responsible for service delivery are referred to as Anganwadi workers. They serve as the main connection between the community and health services, as well as other essential services. Extended mopup rounds were conducted in densely populated urban and peri-urban areas that had consistently shown low coverage in prior IDA rounds. Outreach camps were systematically organized in industrial zones, brick kilns, factories, and transport hubs, including bus stands and railway stations, to engage with individuals who had not yet been reached. The cleanup operation was conducted until 9 March 2019.

### Study Design and Data Gathering

The study evaluated filariasis transmission and control strategies combining field surveys and government health data. Data on IDA coverage, microfilaremia prevalence, and transmission patterns were acquired.

IDA coverage data were collected for three consecutive years (2019, 2020, and 2021) from multiple PHC/CHC regions. For each region, the percentage of people who received triple drug therapy as well as the overall population targeted were noted. By comparing yearly coverage rates and spotting patterns in population participation, the efficacy of IDA was assessed.

Following the WHO guidelines, night blood surveys were carried out between 10:00 p.m. and 2:00 a.m. Participants who lived in endemic areas provided blood samples (2–3 mL). Giemsa stain was applied to thick blood smears, which were then seen under a light microscope. The total number of blood samples collected, examined, and the microfilaria (Mf) rate were recorded for each year.

To evaluate the continuous spread of lymphatic filariasis, the TAS was carried out at random and sentinel sites. A total of 19,852 participants, in which 9,907 from sentinel sites and 9,945 from random sites, were analyzed. Prevalence rates were calculated when microfilaria-positive cases were identified.

### Data Analysis

The percentage of positive cases compared to the total number of samples examined was used to calculate the microfilaria (Mf) rate. The effectiveness of IDA efforts in reducing the spread of disease was assessed by analyzing the yearly trends. To determine the success of intervention strategies, the total Mf rate from sentinel and spot-check sites was evaluated.

## Result

The population coverage for IDA interventions from 2019 to 2021 across the surveyed regions showed a consistent upward trend, reflecting improved implementation and participation. The total population targeted under IDA increased from 4,086,471 in 2019 to 4,154,201 in both 2020 and 2021. Correspondingly, the population coverage improved significantly: **2019**: 3,056,153 individuals were covered, representing **74.79%** coverage. **2020**: Coverage rose to 3,358,375 individuals (**80.84%**), reflecting a 6.05% increase. **2021**: Coverage slightly declined to 3,339,411 individuals (**80.39%**), remaining above 80%. Among individual PHC/CHC regions: The highest percentage of population coverage was consistently observed in **Pindra** (88.79% in 2019, 84.56% in 2020, and 79.76% in 2021). Urban areas demonstrated the lowest coverage throughout the study period, with 65.17% in 2019, 73.79% in 2020, and 79.73% in 2021.Overall, the average IDA coverage across all regions exceeded the 80% threshold in 2020 and 2021, demonstrating significant progress in triple drug therapy.

The microfilaria (Mf) rate across sentinel and spot-check sites showed a marked reduction over the study period, underscoring the impact of IDA interventions. **Sentinel Sites**: In **2019**, a total of 2,017 blood smears (B/S) were examined, yielding an Mf rate of **0.94%** with a total of 19 mf positive individuals in sentinel sites. By **2020**, the Mf rate had reduced to **0.44%**, with 2,008 smears examined. In **2021**, the Mf rate further decreased to **0.15%**, with 2,005 smears examined. Notably, areas such as Bahutara, Badagaon showed complete elimination of Mf (0%) by 2021, while areas such as Kodopur, Ramnagar demonstrated a slight resurgence with a rate of **0.79%** in 2021. **Spot-Check Sites**: The Mf rate at spot-check sites followed a similar declining trend, reducing from **0.34%** in 2020 to **0.05%** in 2021. Sites like Cholapur reported around **1.34%** Mf rate in 2020 while **0.20%** in 2021, while other sites demonstrated sporadic NIL Mf occurrences.

The overall Mf rate (combining sentinel and spot-check sites) decreased significantly over the study period, from **0.46%** in 2019 to **0.39%** in 2020, and finally to **0.10%** in 2021. This reduction reflects the success of the IDA campaigns in implementing lymphatic filariasis transmission.

The Declining Mf rates across all sites indicate effective suppression of disease transmission. The improvement in IDA coverage rates highlights enhanced community participation and intervention reach, particularly in rural areas. The urban regions, although starting with lower coverage, achieved notable progress by 2021, nearly equating rural areas in terms of percentage coverage.

The transmission assessment survey was conducted across multiple sentinel and random sites to evaluate the prevalence of lymphatic filariasis. A total of **19,852** individuals were examined, with **9,907** from sentinel sites and **9,945** from random sites.

Total Positive Cases were Identified 20, Sentinel Sites 6, Random Sites 14 Overall Microfilaria Rate (%) 0.1%, Sentinel Sites 0.06%, Random Sites 0.14%. Sites with positive cases included **Kashi Vidyapeeth (0.33%), Pindra (0.16%)**, **Badagaon (0.16%)**, and **Sevapuri (0.16%)** in rural while sites like **Ramnagar (0.49%)**, **Ashafaq Nagar (0.33%), Anand Mayi (0.33%)** included in urban. Several sites reported zero prevalence, indicating effective transmission control, while the highest prevalence was observed in **Cholapur**, with a microfilaria rate of **1.33%** in rural, ultimately failed the TAS, which requires a threshold of less than 1% for successful intervention of drug therapy. Consequently, additional rounds of IDA is in progress to achieve a successful TAS outcome.

The results indicate **low ongoing transmission** of lymphatic filariasis in the surveyed regions, with a few hotspots requiring targeted interventions. The findings support continued **Triple Drug Therapy** and **vector control measures** in areas with detected microfilaria cases. These results will inform public health strategies to achieve **elimination goals** and sustain the gains made in controlling lymphatic filariasis transmission.

## Discussion

This study introduces the different strategies to determine the LF elimination dataset for three consecutive years from a district, thus representing a substantial contribution to the lymphatic filariasis monitoring in this region. As per the existing research, there is no such longitudinal data that has been reported for this region since last 20 years. The study highlights the significant progress achieved towards the elimination of Lymphatic filariasis (LF) in the Varanasi district surveyed between 2019 to 2021. A substantial decrease in the microfilaria prevalence together with a much lower infection rates as observed under Transmission Assessment Survey results in the beneficial outcomes of the Mass Drug Administration program and enhanced community participation over time.

The sampling strategy, wherein surveys were conducted at high-risk locations assigned based on the prevalence of LF cases, demonstrated ongoing LF transmission in both human and vector populations, with some instances surpassing critical thresholds at the site level (ref). The results show the necessity of ongoing observation, especially in areas where MDA has been conducted or was considered unnecessary. These findings may offer helpful information regarding areas susceptible to revival, particularly in the absence of earlier epidemiological data.

There was a reduction in MF prevalence from 1.7% in 2019 to 0.3% in 2021, indicating the enduring efficacy of the prolonged MDA campaigns. These trends align with the national and worldwide reports demonstrating that sustained high MDA coverage (exceeding 65%) results in major reduction in disease burden (ref).

Similar field-based studies have been documented in many regions of South India, where continuous post-MDA surveillance is guiding public health intervention and policy formation. This study fits with and enhances regional initiatives, underscoring the necessity for localized data to support national elimination objectives. However, in places where MDA and LF control programs are not yet in place, there is always the chance that individuals with residual infection from endemic districts may move to non-MDA districts, leading to transmission in those areas. (ref)

Furthermore, the transmission survey, such as the one provided herein, can provide crucial baseline data for predicting LF elimination by mathematical modelling and simulation. To assess different control scenarios and replicate transmission patterns, these models make use of real-world intervention and epidemiological data, which indicates the ability to forecast when transmission will be interrupted, determines how many rounds of MDA would be necessary, and identifies the region that may have a revival. They could be helpful in national programmatic decisions, and by using this district data to provide elimination.

## Data Availability

All relevant data is within the paper

## Acknowledgements

This study is based upon the MDA/IDA treatment provided by the drug administrators which includes health workers, health vistors, medical officers and locals, we acknowledge their efforts. We are grateful to District Medical officer and all medical and paramedical staff members for night blood test surveys in the population. AS is grateful to Institute of Excellence, BHU for computers, devices and peripherals. Council of science and technology UP. SS is grateful to the University Grant Commission (UGC), New Delhi for providing a Junior Research Fellowship/ Senior Research Fellowship (NTA Ref no. 221610005833). Tripti Singh is thankful to the University Grants Commission (UGC), New Delhi for providing Junior Research Fellowship/ Senior Research Fellowship Ref. No. 211610077377. Sunil Kumar is grateful to the NBCFDC JRF Ref. No. 231620090211 Junior Research Fellowship, University Grants Commission (UGC), New Delhi. Authors are thankful to DBT-BHU Interdisciplinary School of Life Science for providing laboratory space.

## Funding

The study was funded by UP-CST, Lucknow, India through grant (P26/0156)

## Authors’ contributions

SS: Writing original draft, Data curation, Visualization, Methodology analysis, Review and editing; AKS: Designing, Data Curation, Study implementation and Analysis TS: Investigating, Methodology analysis. SK: Formal analysis, Writing, and editing of the manuscript AS: Conceptualization, Data curation, Review and editing, Supervision. All authors read and approved the final manuscript.

## Declaration of competing interest

The authors declare that they have no known competing financial interests or personal relationships that could have appeared to influence the work reported in this paper.

## Tables and Figures

**Table 1.** 50 endemic districts in Uttar Pradesh identified under the lymphatic filariasis elimination program These districts are part of morbidity management and disability prevention and MDA now IDA campaigns to eliminate lymphatic filariasis as a public health problem.

| S.No | Endemic District | S.No | Endemic District | S.No. | Endemic District |
| --- | --- | --- | --- | --- | --- |
| 1 | Allahabad | 18 | Kannauj | 35 | Mau |
| 2 | Ambedgarnagar | 19 | Pratapgarh | 36 | Mirzapur |
| 3 | Amethi | 20 | Bhadoi | 37 | Pilibhit |
| 4 | Auraiya | 21 | Shahjahanpur | 38 | Raibareli |
| 5 | Azamgarh | 22 | Sonbhadra | 39 | Sant Kabir Nagar |
| 6 | Baharaich | 23 | Sultanpur | 40 | Shravasti |
| 7 | Ballia | 24 | Farrukhabad | 41 | Siddharthnagar |
| 8 | Barabanki | 25 | Ghazipur | 42 | Sitapur |
| 9 | Bareilly | 26 | Gorakhpur | 43 | Balrampur |
| 10 | Basti | 27 | Hamirpur | 44 | Banda |
| 11 | Deoria | 28 | Kanpur Dehat | 45 | Chitrakoot |
| 12 | Faizabad | 29 | Kanpur Nagar | 46 | Unnao |
| 13 | Fatehpur | 30 | Kheri | 47 | Varanasi |
| 14 | Gonda | 31 | Kushinagar | 48 | Etawah |
| 15 | Hardoi | 32 | Lucknow | 49 | Chandauli |
| 16 | Jalaun | 33 | Maharajganj | 50 | Kaushambi |
| 17 | Jaunpur | 34 | Mahoba |  |  |

**Table 2.** IDA coverage over different PHC/CHC for 2019-2021 The percentage of population covered under IDA in eight PHC/CHC of Varanasi including urban areas for the year 2019,2020 and 2021. Steady progress or fluctuation can be observed across the years, reflecting efforts in improving drug administration.

| S.N | Name of PHC/CHC | 2019 |  |  | 2020 |  |  | 2021 |  |  |
| --- | --- | --- | --- | --- | --- | --- | --- | --- | --- | --- |
|  |  | Total Population | Population Covered in IDA | % Population covered in IDA | Total Population | Population Covered in IDA | % Population covered in IDA | Total Population | Population Covered in IDA | % Population covered in IDA |
| 1 | HARAHUVA | 253626 | 211545 | 83.40824679 | 267442 | 238552 | 89.19765781 | 267442 | 222580 | 83.2255218 |
| 2 | SEVAPURI | 240184 | 200114 | 83.316957 | 260615 | 219620 | 84.26990004 | 260615 | 216749 | 83.16827504 |
| 3 | KASHI VIDYA PEETH | 302227 | 259343 | 85.81066549 | 384574 | 319631 | 83.11300296 | 384574 | 315505 | 82.04012752 |
| 4 | ARAZILINE | 365704 | 292410 | 79.9581082 | 365704 | 320105 | 87.53117275 | 365704 | 292653 | 80.02455538 |
| 5 | BADAGAON | 229908 | 187481 | 81.5460967 | 232759 | 195881 | 84.15614434 | 232759 | 185389 | 79.64847761 |
| 6 | CHIRAIGAON | 291995 | 247208 | 84.66172366 | 305000 | 268763 | 88.11901639 | 332000 | 266133 | 80.16054217 |
| 7 | CHOLAPUR | 299735 | 221228 | 73.80786361 | 281996 | 247059 | 87.61081717 | 281996 | 222549 | 78.91920453 |
| 8 | PINDRA | 279950 | 248555 | 88.78549741 | 292325 | 247206 | 84.56546652 | 292325 | 233160 | 79.76054049 |
| 9 | URBAN | 1823142 | 1188269 | 65.17698567 | 1763786 | 1301558 | 73.79341938 | 1736786 | 1384693 | 79.72732392 |
| 10 | TOTAL | 4086471 | 3056153 | 74.78709625 | 4154201 | 3358375 | 80.84286244 | 4154201 | 3339411 | 80.3863607 |

**Table 3.**
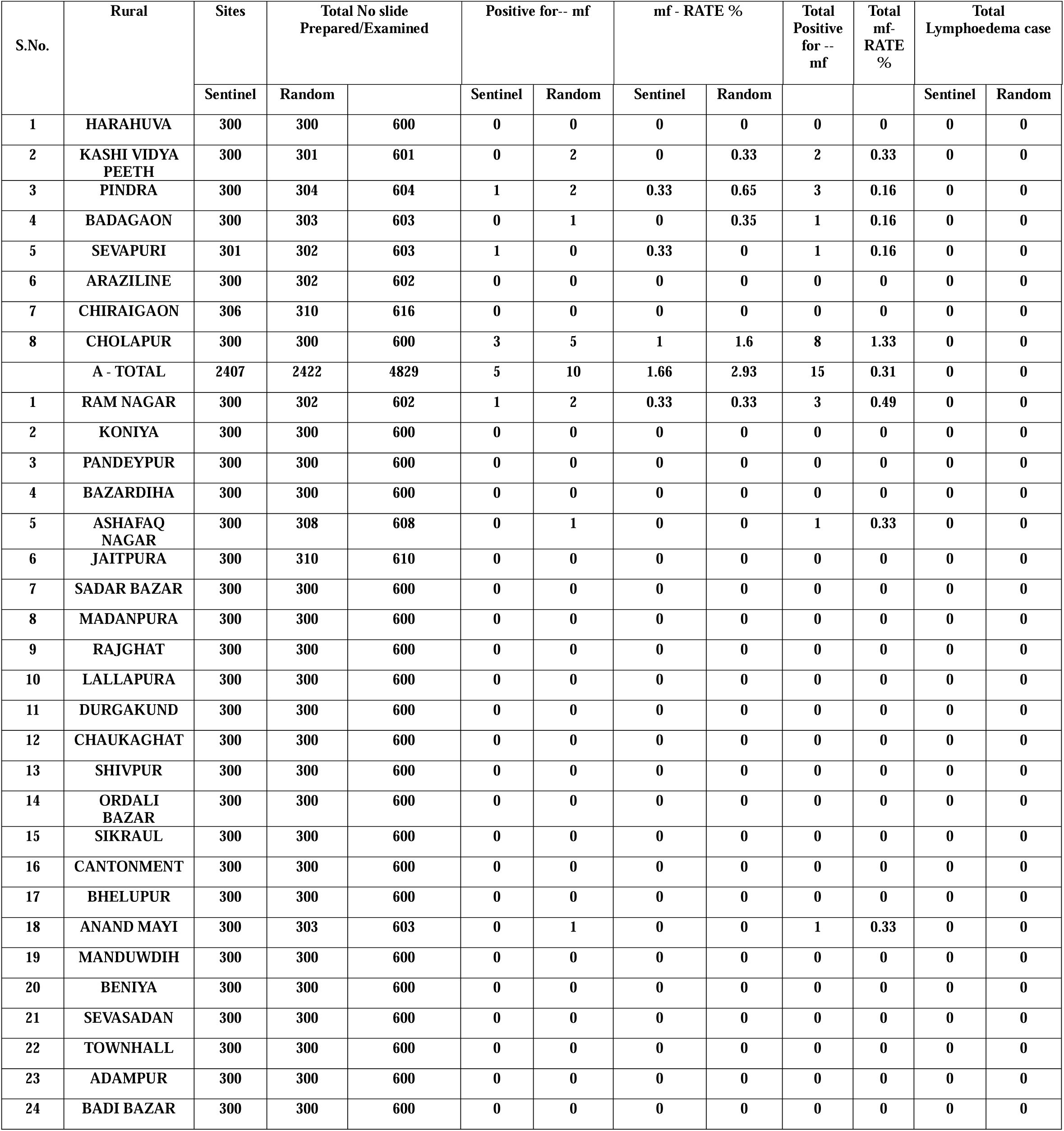
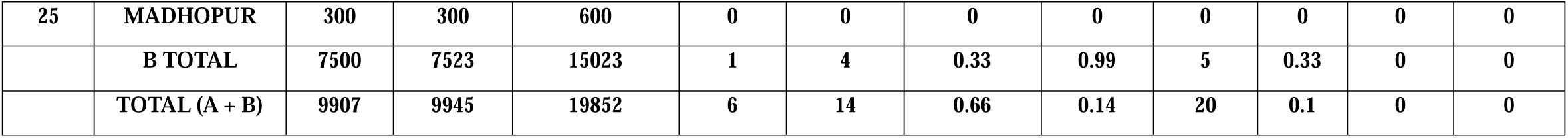
TAS conducted across various sentinel and random sites under the lymphatic filariasis elimination program Result of TAS conducted across various sentinel and random sites under the lymphatic filariasis elimination programme

| S.No. | Rural | Sites | Total No slide Prepared/Examined |  | Positive for-- mf |  | mf - RATE % |  | Total Positive for -- mf | Total mf-RATE % | Total Lymphoedema case |  |
| --- | --- | --- | --- | --- | --- | --- | --- | --- | --- | --- | --- | --- |
|  |  | Sentinel | Random |  | Sentinel | Random | Sentinel | Random |  |  | Sentinel | Random |
| 1 | HARAHUVA | 300 | 300 | 600 | 0 | 0 | 0 | 0 | 0 | 0 | 0 | 0 |
| 2 | KASHI VIDYA PEETH | 300 | 301 | 601 | 0 | 2 | 0 | 0.33 | 2 | 0.33 | 0 | 0 |
| 3 | PINDRA | 300 | 304 | 604 | 1 | 2 | 0.33 | 0.65 | 3 | 0.16 | 0 | 0 |
| 4 | BADAGAON | 300 | 303 | 603 | 0 | 1 | 0 | 0.35 | 1 | 0.16 | 0 | 0 |
| 5 | SEVAPURI | 301 | 302 | 603 | 1 | 0 | 0.33 | 0 | 1 | 0.16 | 0 | 0 |
| 6 | ARAZILINE | 300 | 302 | 602 | 0 | 0 | 0 | 0 | 0 | 0 | 0 | 0 |
| 7 | CHIRAI GAON | 306 | 310 | 616 | 0 | 0 | 0 | 0 | 0 | 0 | 0 | 0 |
| 8 | CHOLAPUR | 300 | 300 | 600 | 3 | 5 | 1 | 1.6 | 8 | 1.33 | 0 | 0 |
|  | A - TOTAL | 2407 | 2422 | 4829 | 5 | 10 | 1.66 | 2.93 | 15 | 0.31 | 0 | 0 |
| 1 | RAM NAGAR | 300 | 302 | 602 | 1 | 2 | 0.33 | 0.33 | 3 | 0.49 | 0 | 0 |
| 2 | KONIYA | 300 | 300 | 600 | 0 | 0 | 0 | 0 | 0 | 0 | 0 | 0 |
| 3 | PANDEYPUR | 300 | 300 | 600 | 0 | 0 | 0 | 0 | 0 | 0 | 0 | 0 |
| 4 | BAZARDIHA | 300 | 300 | 600 | 0 | 0 | 0 | 0 | 0 | 0 | 0 | 0 |
| 5 | ASHAFAQ NAGAR | 300 | 308 | 608 | 0 | 1 | 0 | 0 | 1 | 0.33 | 0 | 0 |
| 6 | JAITPURA | 300 | 310 | 610 | 0 | 0 | 0 | 0 | 0 | 0 | 0 | 0 |
| 7 | SADAR BAZAR | 300 | 300 | 600 | 0 | 0 | 0 | 0 | 0 | 0 | 0 | 0 |
| 8 | MADANPURA | 300 | 300 | 600 | 0 | 0 | 0 | 0 | 0 | 0 | 0 | 0 |
| 9 | RAJGHAT | 300 | 300 | 600 | 0 | 0 | 0 | 0 | 0 | 0 | 0 | 0 |
| 10 | LALLAPURA | 300 | 300 | 600 | 0 | 0 | 0 | 0 | 0 | 0 | 0 | 0 |
| 11 | DURGAKUND | 300 | 300 | 600 | 0 | 0 | 0 | 0 | 0 | 0 | 0 | 0 |
| 12 | CHAUKAGHAT | 300 | 300 | 600 | 0 | 0 | 0 | 0 | 0 | 0 | 0 | 0 |
| 13 | SHIVPUR | 300 | 300 | 600 | 0 | 0 | 0 | 0 | 0 | 0 | 0 | 0 |
| 14 | ORDALI BAZAR | 300 | 300 | 600 | 0 | 0 | 0 | 0 | 0 | 0 | 0 | 0 |
| 15 | SIKRAUL | 300 | 300 | 600 | 0 | 0 | 0 | 0 | 0 | 0 | 0 | 0 |
| 16 | CANTONMENT | 300 | 300 | 600 | 0 | 0 | 0 | 0 | 0 | 0 | 0 | 0 |
| 17 | BHELUPUR | 300 | 300 | 600 | 0 | 0 | 0 | 0 | 0 | 0 | 0 | 0 |
| 18 | ANAND MAYI | 300 | 303 | 603 | 0 | 1 | 0 | 0 | 1 | 0.33 | 0 | 0 |
| 19 | MANDUWDIH | 300 | 300 | 600 | 0 | 0 | 0 | 0 | 0 | 0 | 0 | 0 |
| 20 | BENIYA | 300 | 300 | 600 | 0 | 0 | 0 | 0 | 0 | 0 | 0 | 0 |
| 21 | SEVASADAN | 300 | 300 | 600 | 0 | 0 | 0 | 0 | 0 | 0 | 0 | 0 |
| 22 | TOWNHALL | 300 | 300 | 600 | 0 | 0 | 0 | 0 | 0 | 0 | 0 | 0 |
| 23 | ADAMPUR | 300 | 300 | 600 | 0 | 0 | 0 | 0 | 0 | 0 | 0 | 0 |
| 24 | BADI BAZAR | 300 | 300 | 600 | 0 | 0 | 0 | 0 | 0 | 0 | 0 | 0 |
| 25 | MADHOPUR | 300 | 300 | 600 | 0 | 0 | 0 | 0 | 0 | 0 | 0 | 0 |
|  | B TOTAL | 7500 | 7523 | 15023 | 1 | 4 | 0.33 | 0.99 | 5 | 0.33 | 0 | 0 |
|  | TOTAL (A + B) | 9907 | 9945 | 19852 | 6 | 14 | 0.66 | 0.14 | 20 | 0.1 | 0 | 0 |

**Table 4.** NBT survey analysis for sentinel and spot check regions (2019-2021) NBT result across sentinel and spot check sites over three years. It includes data on the no. of blood smears collected and examined along with microfilariae. There is marked declined in mf rates over the years with most regions reporting NIL in 2020 and 2021 reflecting progress in disease management efforts.

| S. No. | Particulars | 2019 |  |  |  |  | 2020 |  |  |  |  | 2021 |  |  |  |  |
| --- | --- | --- | --- | --- | --- | --- | --- | --- | --- | --- | --- | --- | --- | --- | --- | --- |
|  |  | Name of the sites<br>CHC/PHC,<br>VILLAGE | TOTAL<br>B/S<br>Collection | Total B/S<br>Examined | No.<br>of<br>+ve<br>for<br>Mf | Mf<br>rate<br>(%) | Name of the sites<br>CHC/PHC/SUB-CENTER/WARD<br>VILLAGE | TOTAL<br>B/S<br>Collection | Total B/S<br>Examined | No.<br>of<br>+ve<br>for<br>Mf | Mf<br>rate<br>(%) | Name of<br>SitesCHC/PHC | Total B/S<br>Collected | Total B/S<br>Examined | No.<br>of<br>+ve<br>for<br>Mf | Mf rat<br>(%) |
| 1 | Sentinel<br>(Rural) | Bahutara<br>Badagaon | 500 | 500 | 7 | 1.40% | Badagaon<br>Basani<br>Bahutara | 503 | 503 | NIL | NIL | Babutara<br>Badagaon | 500 | 500 | NIL | NIL |
| 2 | Sentinel<br>(Rural) | Bhullanpur<br>Kashi Vidya<br>Peeth | 501 | 501 | 6 | 1.20% | Pindra<br>Jamapur<br>Jamapur | 502 | 502 | 4 | 0.79% | Jamapur<br>Pindra | 500 | 500 | 2 | 0.40% |
| 3 | Sentinel<br>(Rural) | Jamapur<br>Pindra | 500 | 500 | 3 | 0.60% | Kashi Vidya<br>Peeth Govindpur<br>Bhullanpur | 501 | 501 | 1 | 0.19% | Bhullanpur<br>Kashi<br>Vidyapeeth | 500 | 500 | NIL | NIL |
| 4 | Sentinel<br>(Urban) | Kodopur<br>Ramnagar | 516 | 516 | 3 | 0.58% | Ramnagar<br>Ward No-18<br>Tapovan | 502 | 502 | 4 | 0.79% | Kutalupur,<br>Rattapur<br>Ramnagar | 505 | 505 | 1 | 0.20% |
| Sentinel Sites Subtotal (A) |  |  | 2017 | 2017 | 19 | 0.94% |  | 2008 | 2008 | 9 | 0.44% |  | 2005 | 2005 | 3 | 0.15% |
| 1 | Spot<br>Check<br>(Rural) | Raunakhurd<br>Cholapur | 503 | 503 | NIL | NIL | Cholapur<br>Munari<br>Munari | 520 | 520 | 7 | 1.34% | Jagdeeshpur<br>Cholapur | 500 | 500 | 1 | 0.20% |
| 2 | Spot<br>Check<br>(Rural) | Ganeshpur<br>Araziline | 504 | 504 | NIL | NIL | Sevapuri<br>Kardhana<br>Kardhana | 507 | 507 | NIL | NIL | Matuka<br>Sevapuri | 500 | 500 | NIL | NIL |
| 3 | Spot<br>Check<br>(Rural) | Behra<br>Sevapuri | 556 | 556 | NIL | NIL | Chiragaon<br>Barai<br>Barai | 506 | 506 | NIL | NIL | Baral<br>Chiragaon | 502 | 502 | NIL | NIL |
| 4 | Spot<br>Check<br>(Urban) | Nagwan<br>Aandmayi | 505 | 505 | NIL | NIL | Orderly Bazar<br>Thithori Mahal<br>Ordely Bazar | 500 | 500 | NIL | NIL | Bhojuveer<br>Shivpur | 505 | 505 | NIL | NIL |
| Random Sites Sub Total (B) |  |  | 2068 | 2068 | NIL | NIL |  | 2033 | 2033 | 7 | 0.34 |  | 2007 | 2007 | 1 | 0.05% |
| TOTAL (A + B) |  |  | 4085 | 4085 | 19 | 0.46% |  | 4041 | 4041 | 16 | 0.39% |  | 4012 | 4012 | 4 | 0.10% |

**Figure 1.**
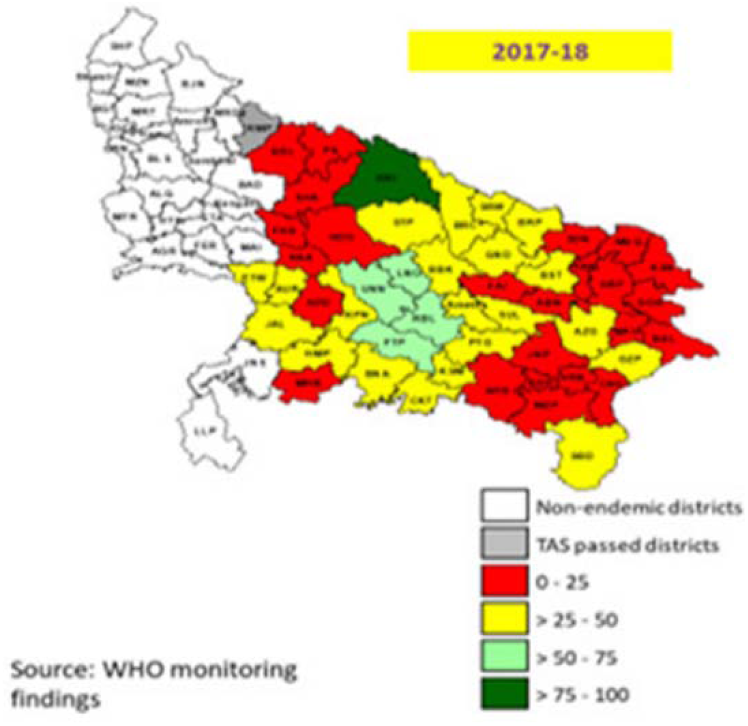
Geographical distribution of lymphatic filariasis endemicity in Uttar Pradesh for 2017-18 The map shows the distribution of endemic districts in Uttar Pradesh based on WHO monitoring data. Districts are categorised by TAS status and the percentage coverage achieved: non endemic districts (white), TAS passed district (grey), endemic districts with varying coverage levels (red 0-25%, orange 25-50%, yellow 50-75%, green 75-100%). The map highlights progress in MDA coverage and areas requiring intensified efforts for elimination.

**Figure 2.**
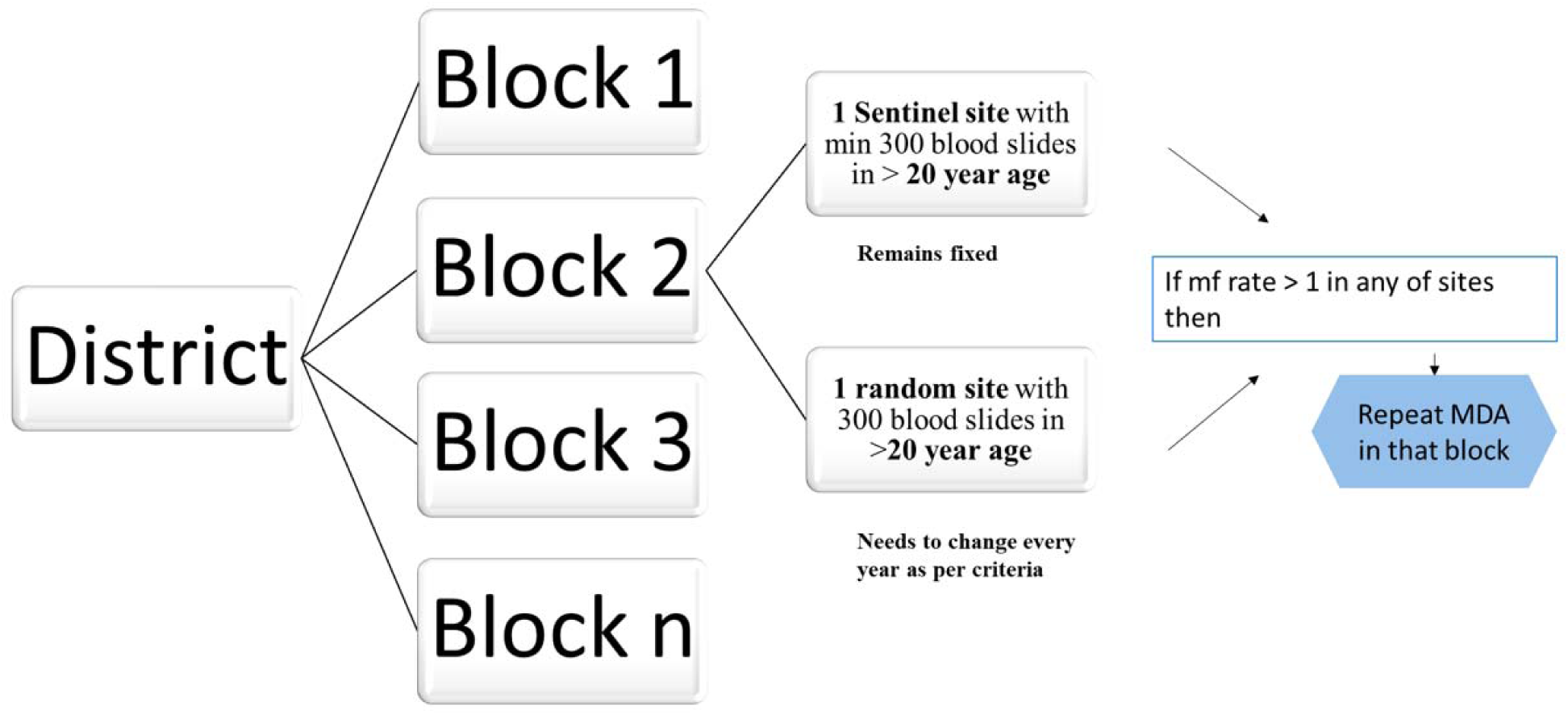
Microfilariae assessment survey framework at the block level The flow diagram outlines the framework for conducting the microfilariae assessment survey at the block level to evaluate the elimination of lymphatic filariasis. Each district is divided into multiple blocks where the survey is conducted.

**Figure 3.**
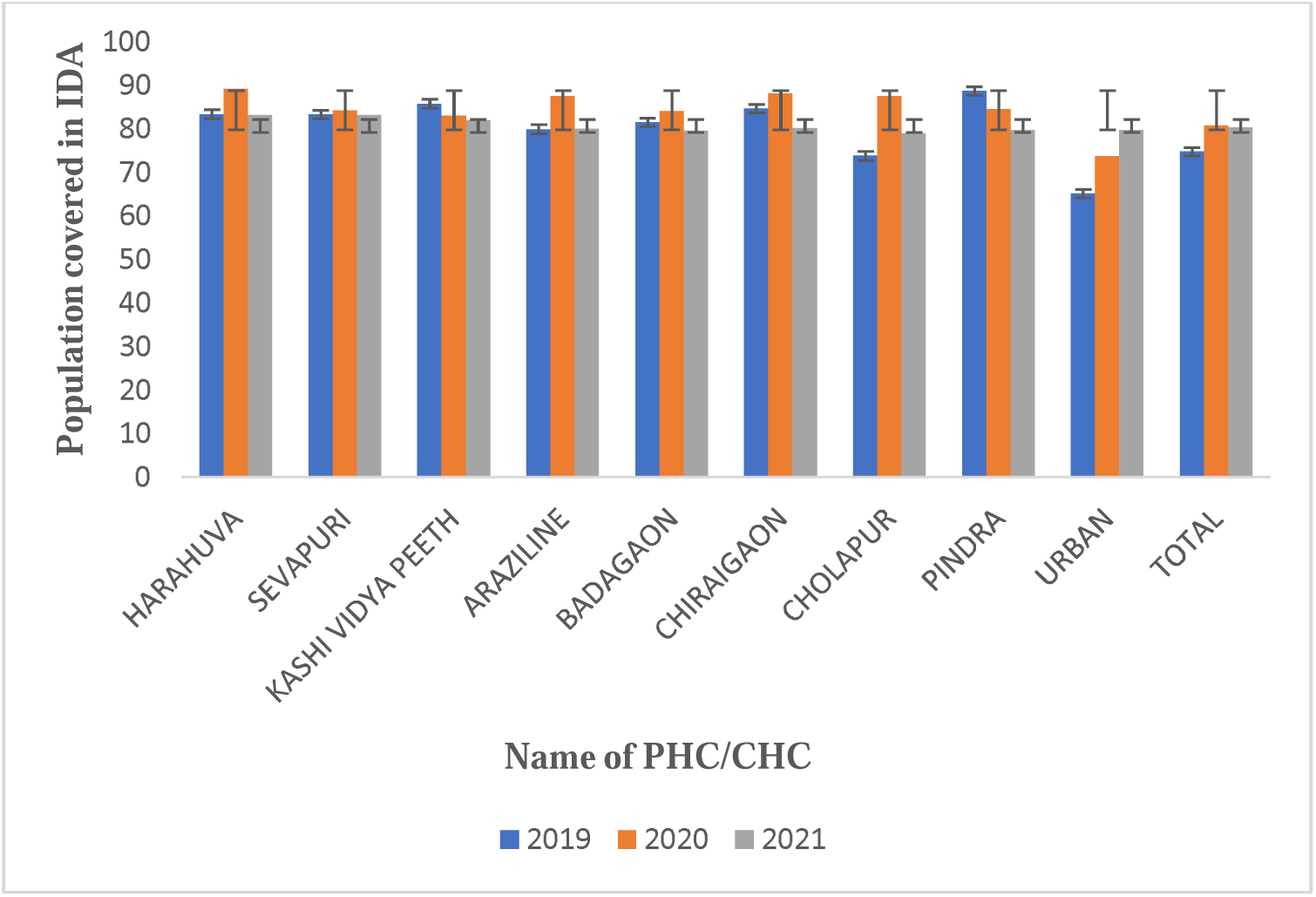
The percentage of the population covered under IDA in eight PHC/CHC of Varanasi including urban areas for 2019-2021. The percentage of population covered under IDA in eight PHC/CHC of Varanasi including urban areas for the year 2019 to 2021 and show the coverage exceeding 65% in most of the areas.

**Figure 4.**
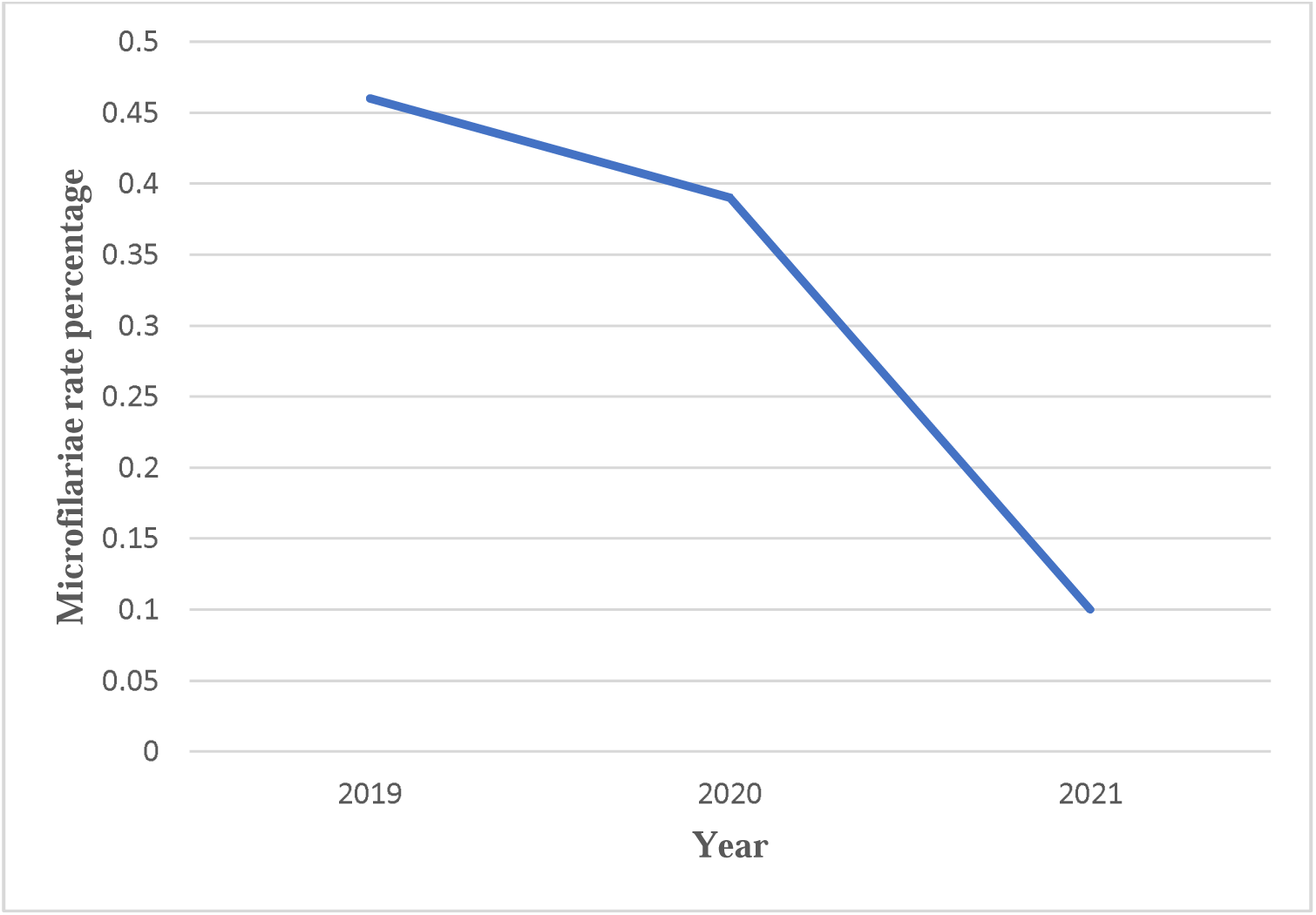
Microfilariae rate percent from year 2019-2021 The graph shows the percentage of microfilariae rates recorded annually from 2019 to 2021 demonstrating a consistent decline over the three periods. This decline suggests the effectiveness of IDA/MDA strategies implemented during this period.

**Figure 5.**
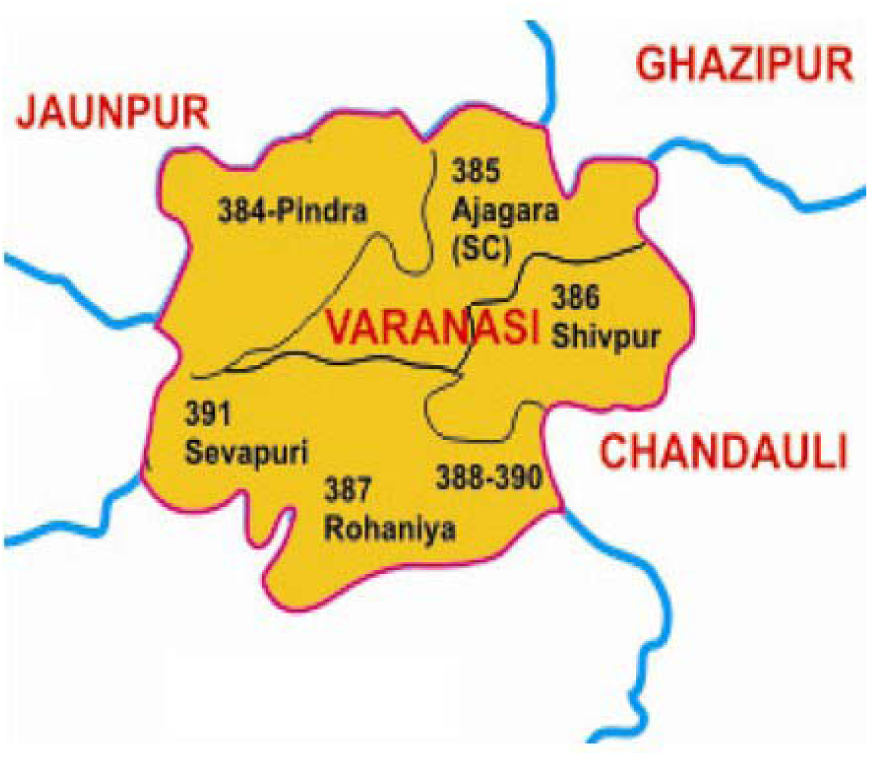
Workflow for Lymphatic Filariasis Surveillance and Elimination Strategy This flow diagram depicts a field-based approach to control and eliminate lymphatic filariasis. The process begins with planning and preparation strategies, which include defining study areas, mapping them, and training personnel. The next step is to gather the field data, which involves the engagement of the community, collecting surveys and blood samples, and eventually detecting the microfilariae. The next step involves data entry, analysis, and reporting of the additional data that has been collected. Mass drug administration (MDA) and continuous evaluation are the intervention and monitoring actions that are begun based on the findings. In this last phase, we focus on the public health consequences, namely how the Filaria Mukt Abhiyan is working to control the disease and eventually eradicate it.

**Figure 6.**
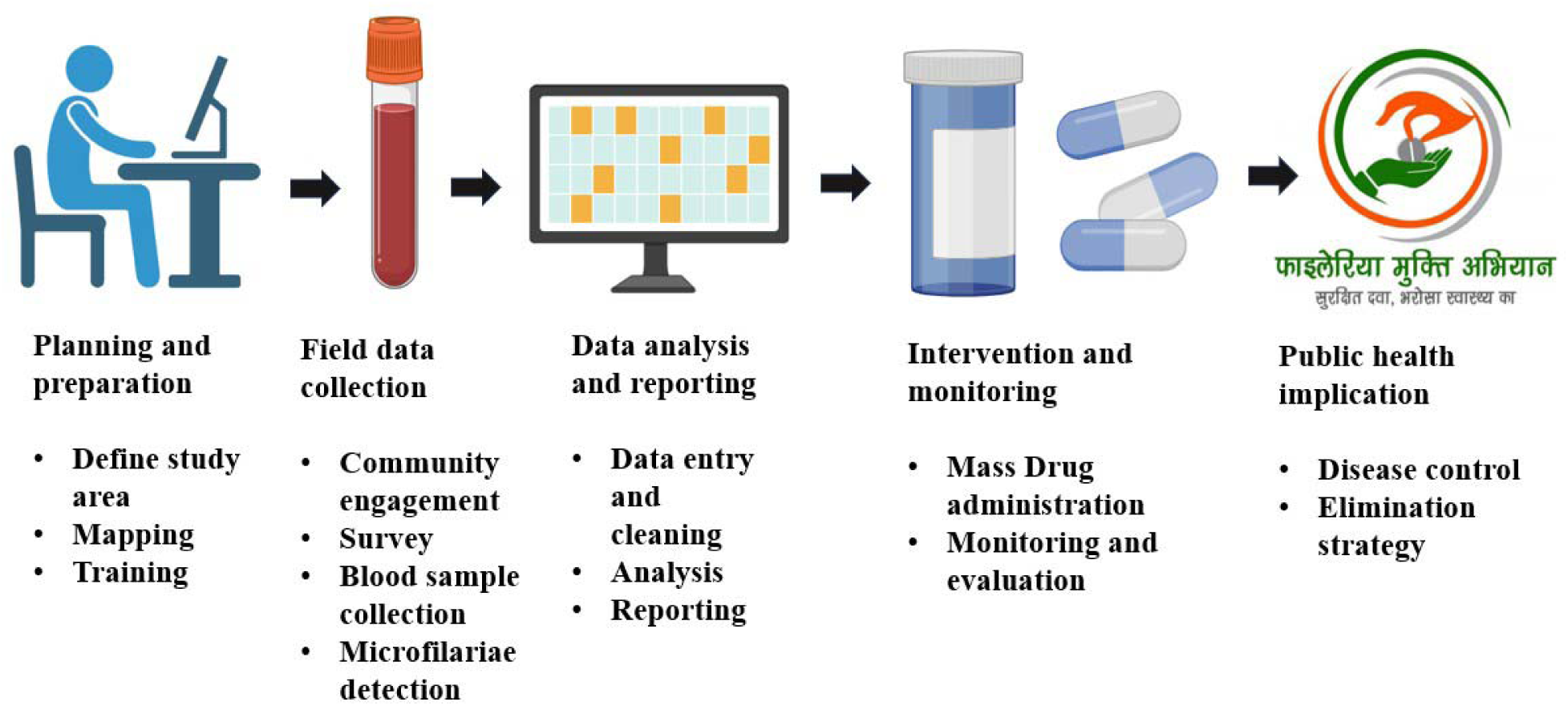
Workflow for Lymphatic Filariasis Surveillance and Elimination Strategy

## Notes

### Competing Interest Statement

The authors have declared no competing interest.

## References

1. de Souza, D. K., K. Gass, J. Otchere, Y. M. Htet, O. Asiedu, B. Marfo, N.-K. Biritwum, D. A. Boakye and C. S. Ahorlu (2020). “Review of MDA registers for lymphatic filariasis: findings, and potential uses in addressing the endgame elimination challenges.” PLoS Neglected Tropical Diseases 14(5): e0008306.

2. Parveen, K. and L. D. Singh (2023). “Evaluation of Triple Drug Administration for Lymphatic Filariasis in Prayagraj District, Uttar Pradesh, India: A Cross-sectional Study.” Journal of Clinical & Diagnostic Research 17(6).

3. Rebollo, M. P. and M. J. Bockarie (2013). “Toward the elimination of lymphatic filariasis by 2020: treatment update and impact assessment for the endgame.” Expert review of anti-infective therapy 11(7): 723–731.

4. Ton, T. G., C. Mackenzie and D. H. Molyneux (2015). “The burden of mental health in lymphatic filariasis.” Infectious diseases of poverty 4(1): 34.

5. Tripathi, B., N. Roy and N. Dhingra (2022). “Introduction of triple-drug therapy for accelerating lymphatic filariasis elimination in India: lessons learned.” The American Journal of Tropical Medicine and Hygiene 106(5 Suppl): 29.

6. WHO (2010). Global Programme to Eliminate Lymphatic Filariasis: Progress Report 2000–2009 and Strategic Plan 2010–2020, World Health Organization Geneva.

7. Wynd, S., W. D. Melrose, D. N. Durrheim, J. Carron and M. Gyapong (2007). “Understanding the community impact of lymphatic filariasis: a review of the sociocultural literature.” Bulletin of the World Health Organization 85: 493–498.

